# Factors associated with support-seeking behavior among AIAN women survivors of IPV

**DOI:** 10.1101/2022.12.06.22283189

**Authors:** Susanna Y Park

**Author notes:** Corresponding author: (SP). 1890 N Revere Ct, Aurora, CO, USA.

## Abstract

**Objectives:** To understand how Native women seek support as survivors of intimate partner violence by examining factors associated with seeking formal support (e.g., medical, police), informal support (e.g., family, friends), or both types of support.

**Methods:** Data from the 2010 National Intimate and Sexual Violence Survey were analyzed among the AIAN oversamples. Descriptive statistics and odds ratio analyses were conducted to observe significant factors that are associated with adult Native women survivors seeking support.

**Results:** Survivors have higher odds of seeking informal supports than formal supports. Formal support is not often sought, however survivors had significantly higher odds of seeking therapy. Conclusions. While there are various avenues in which survivors may seek support, not all types of support are helpful to the survivor’s needs. However, this study is still valuable in distinguishing factors related to support-seeking and observing general relationships. Focused and intentional public health research on support for AIAN women survivors is needed to further understand how programs and policies can be improved.

## Introduction

Intimate partner violence (IPV) is the most common type of violence experienced by American Indian/Alaska Native (AIAN) women and is a significant public health issue. [1–4].^2^ A 2016 report from the National Intimate Partner and Sexual Violence Survey (NISVS) revealed that more than four in five (84.3%) AIAN adult (ages 18-64) women have experienced violence in their lifetime and more than one in three AIAN adult women experienced violence in the past year. Of these women, 14.4% experienced sexual violence, 8.6% experience physical violence by an intimate partner, 25.5% experienced psychological aggression by an intimate partner, and 11.6% experienced stalking [4]. AIAN women are at a significantly higher risk of experiencing stalking and physical violence by an intimate partner in their lifetime than their non-Hispanic White-only counterparts [1,2,5]. In addition, they are significantly more likely to experience psychological aggression by an intimate partner in their lifetime in the past year [4]. Such experiences of violence severely impact AIAN women’s physical, mental, emotional, and spiritual well-being [5,6]. For the purpose of this research, IPV refers to four types of behaviors perpetrated by a current or former spouse, and dating partners: physical violence, sexual violence, stalking, and psychological aggression [7].

## Support seeking

Effective and relevant services are needed for AIAN women survivors of violence. According to findings from the NISVS, among AIAN adult women surveyed, 38% of women needed medical care, 16% needed legal services, 11% needed housing services, and 9% needed advocacy services. However, 38% of AIAN women were unable to get the services they needed. Further, they face various barriers to seeking needed services. AIAN women were found to be 2.5 times as likely as non-Hispanic White-only women to lack access to needed services [4].

While interventions that focus on positive social support may benefit victims, the magnitude of the impact is small [8]. Individual barriers include factors that deter the woman from seeking support due to her perceptions regarding the resources available to her. Systemic barriers include factors outside of the individual’s control that may prevent the woman from seeking support. AIAN women are unlikely to report their experiences to the police due to fear of being judged, fear of not being believed, fear of being stereotyped as a Native person, and fear that authorities will not understand [9]. Women may also turn away from seeking support in fear of being blamed and stigmatized for their experience [10]. The added layer of Native identity is often difficult to navigate in settings where AIAN women are more likely to experience discrimination based on their ethnicity.

Numerous studies on the support-seeking behaviors among survivors of IPV indicate various factors that may encourage victims to seek support. Fleming & Resick (2017) reported that age, depression, psychological aggression, PTSD, perceived helpfulness of resource, and perceived controllability of violence were significantly related to support-seeking behavior. High levels of education and employment are found to be positively associated with support-seeking behaviors [11,12]. Women who also have more autonomy and decision-making agency may increase their likelihood of seeking support. Conversely, more autonomy and decision making can also lead to women not seeking support due to fear of what may happen once they do seek support [12]. If the woman has a child, her decision to seek support is further influenced by seeking options that meet the best interests of the child (such as seeking alternative housing options). This indicates a need for policies and programs that protect both the mother and child [13].

Survivors are more likely to seek informal support (e.g., friends and family) than seek formal support (e.g., police or medical professional) [11,14,15]. When the violence is severe, women are more likely to seek either informal or formal support [11,12,16]. However, if women perceive the violence as justified or engage in self-blaming behavior, they may be less likely to seek support in general [11].

Informal support may lead to seeking formal support. For instance, domestic violence survivors who sought informal support also decided to seek formal support if the family or friend they disclosed to had experienced or had knowledge of domestic violence [17]. For Native women, informal care is often protective if the family or social network is tight-knit, supportive, and affirming. However, if there is a risk of breaching confidentiality and lack of accountability [18], women are less likely to access informal network for support and may seek formal support [16].

The intent of this study is to understand how Native women seek support as survivors of intimate partner violence. The 2010 National Intimate and Sexual Violence Survey is the most recent data available, and the AIAN oversample was analyzed. Descriptive statistics and odds ratio analyses were conducted to observe significant factors associated with survivors seeking support.

## Methods

### Data

Secondary data analysis was conducted using data from the 2010 National Intimate and Sexual Violence Survey (NISVS). The survey is maintained by the Centers for Disease Control and Prevention (CDC) and provides the most recent, nationally representative data on violence against woman. The NISVS is the most comprehensive on-going survey that contains national- and state-level data on intimate partner violence, sexual violence, and stalking victimization in the United States [7]. This dataset contains an oversample of AIAN individuals on the various experiences of violence, perpetrator-level data, and the support-seeking behavior among victims of violence. The AIAN oversample raw data are what this project utilized. From January 22 to December 31, 2010, sampling of the AIAN population was maximized via landline telephone exchanges where at least 50% of the population was estimated to be AIAN, which included a total of 16 states in North America.

### Ethics statement

The study was approved by Oregon State University’s Institutional Review Board (IRB-2019-0409). The dataset was obtained with IRB approval through ICPSR data repository (ICPSR 36140).

### Statistical analysis

The study sample used in the empirical analysis was limited to AIAN women who reported having experienced IPV in their lifetime and were between the ages 18-64 at the time of survey (N=1275). A logistic regression model was utilized as follows:

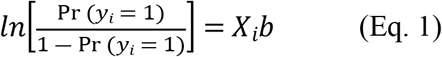

where *yi* refers to support-seeking behavior (support only, informal support only, or both formal and informal supports) for individual *i*, and e is the idiosyncratic error term. “*Xi*” include all factors that may be associated with support-seeking behavior at individual, relational, community, and systemic levels. “b” is a vector of coefficients. Several sensitivity and specification analyses were performed. A linear probability model (LPM) via ordinary least squares (OLS) was estimated as this estimation technique renders consistent and unbiased parameter estimates for binary outcomes.

Descriptive statistics are reported without weights due to the high number of missing observations with weights applied. Data analysis was completed via Stata 15, a statistical software package for data management and analysis [19].

## Results

Descriptive statistics (Table 1) reveal that among AIAN women between 18 to 64 years old who experienced IPV in their lifetime (N=1275), 55.9% sought formal support only, 77.4% sought informal support only, and 50.8% sought both formal and informal types of support. The average age is 41 years old. Of this sample, 80.8% are enrolled in a tribe. 80.7% of the respondents report ever having a child live with them. Few report being gay or lesbian (0.6%), or bisexual (3.0%), and 87.1% identify as heterosexual. 36.6% report as being married, 34.8% report being never married and 20.5% report being divorced, separated, or widowed. Perceived physical and mental health statuses are indicated on a scale of one to five, with one being the worst and five being the best. The mean perceived health status is 3.1 and the perceived mental health status is 3.5.

**Table 1.** Variables, Definitions, and Summary Statistics

| Variables | Frequency (N=1275) | Sample mean or % |
| --- | --- | --- |
| *Dependent Variables* |  |  |
| Formal support only | 713 | 55.9% |
| Informal support only | 987 | 77.4% |
| Formal and Informal support | 647 | 50.8% |
| *Demographic* |  |  |
| Age 18-64yrs (SD) |  | 41 (13.3) |
| Enrolled in tribe | 1021 | 80.8% |
| Ever had child live with you | 1028 | 80.7% |
| *Sexual Orientation* |  |  |
| Heterosexual* | 1110 | 87.1% |
| Gay/lesbian | 7 | 0.6% |
| Bisexual | 38 | 3.0% |
| *Household Income* |  |  |
| Low (< \$20k) | 516 | 40.5% |
| Middle (\$20k to < \$50k) | 1001 | 78.5% |
| High (\$50k+)* | 235 | 18.4% |
| *Marital Status* |  |  |
| Married* | 467 | 36.6% |
| Divorced, separated, or widowed | 267 | 20.5% |
| Never married | 444 | 34.8% |
| *Education level* |  |  |
| Primary education or some high school* | 197 | 15.5% |
| High school diploma | 369 | 28.9% |
| Vocational school | 58 | 4.6% |
| Some college | 414 | 32.5% |
| College diploma or above | 235 | 18.4% |
| *Health status* |  |  |
| Perceived physical health status (1-5)<br>(SD) |  | 3.1 (1.1) |
| Perceived mental health status (1-5)<br>(SD) |  | 3.5 (1.1) |
| *Services sought and quality (=1 if helpful)* |  |  |
| Police | 538 | 45.1% |
| Police quality | 412 | 76.7% |
| Medical | 319 | 26.7% |
| Medical quality | 277 | 87.7% |
| Therapy | 419 | 35.2% |
| Therapy quality | 389 | 93.3% |
| Family | 742 | 62.2% |
| Family quality | 653 | 93.5% |
| Intimate partner | 342 | 28.7% |
| Intimate partner quality | 224 | 93.7% |
| Friend | 820 | 68.7% |
| Friend quality | 787 | 96.8% |

*\*Reference variable*

Household income was divided into three ranges based on the median income among AIANs [20]. 40.5% of respondents are in the low-income range of less than $20k, 78.5% are in the middle-income range of $20k to less than $50k, and 18.4% are in the high-income range of earning $50k or more. Educational attainment was divided into five categories: primary education or some high school (15.5%), high school diploma (28.9%), vocational school (4.6%), some college (32.5%), and college diploma or above (18.4%).

Specific types of formal (e.g., police, medical, therapy) and informal (family, friend, intimate partner) supports were examined for the percentage of respondents who sought a specific type of support, and if they found it to be helpful. A service was considered helpful in this instance if the respondent indicated that the service was a little, somewhat, or very helpful. For instance, 45.1% of respondents sought police support, and of these respondents 76.7% indicated that it was helpful. 26.7% of respondents sought medical support and 87.7% indicated that it was helpful. Less than half of the respondents sought therapy (35.2%) but 93.3% indicated that it was helpful. Among those who sought informal support 62.2% reached out to family, and 93.5% reported it was helpful. 68.7% sought informal support from a friend, and 96.8% of those respondents reported that it was helpful. Friends may include friends, coworkers, acquaintances, or other non-family and friend relationships. Lastly, 28.7% sought support from an intimate partner, and 93.7% of those reported that it was helpful.

### Formal, informal, and both types of support

In this first stage of analysis (Table 2), formal support included medical, legal services, community advocacy, and housing services. Informal support included family, friends, and intimate partners. Adjusted odds ratios were calculated using logistic regression analysis with robust standard errors for adult AIAN women survivors who experienced IPV in their lifetime. Compared to those who are married, those who are divorced, separated, or widowed have higher odds of seeking formal support (adjusted OR=2.23, 95% CI=1.61 to 3.09) and seeking both formal and informal supports (OR=1.89, 95% CI= 1.39 to 2.58). Compared to those who never had a child live with them, the odds of seeking informal support only are 1.46 times (95% CI=1.05 to 2.04) as high for women who ever had a child live with them. The odds of seeking both types of support for women who ever had a child live with them is 1.39 times (95% CI= 1.03 to 1.87) as high. Compared to adult AIAN women survivors with high household income, those with low household income have lower odds of seeking informal support (OR=0.65, 95% CI=0.47 to 0.90). Those who live with middle household income are 1.37 times (95% CI=1.00 to 1.88) as likely to seek both formal and informal types of support. Compared to those who completed primary education or attended some high school, women who attended vocational school have higher odds of seeking informal support (OR=2.59, 95% CI=1.09 to 6.09). Those who have a college diploma or above also have higher odds of seeking informal support (OR=1.73, 95% CI=1.03 to 2.89).

**Table 2.**
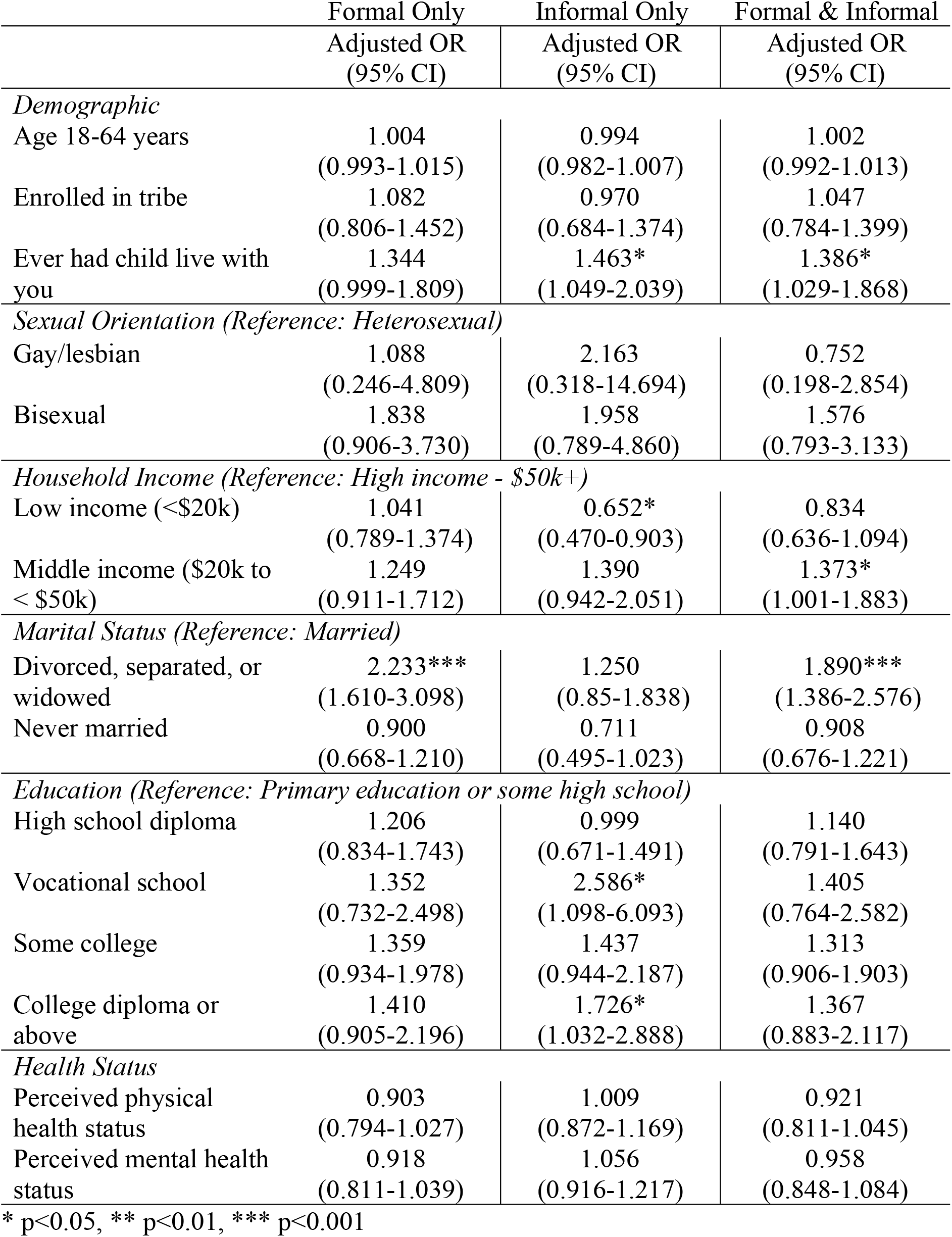
Adjusted ORs for types of support

|  | Formal Only | Informal Only | Formal & Informal |
| --- | --- | --- | --- |
|  | Adjusted OR<br>(95% CI) | Adjusted OR<br>(95% CI) | Adjusted OR<br>(95% CI) |
| *Demographic* |  |  |  |
| Age 18-64 years | 1.004<br>(0.993-1.015) | 0.994<br>(0.982-1.007) | 1.002<br>(0.992-1.013) |
| Enrolled in tribe | 1.082<br>(0.806-1.452) | 0.970<br>(0.684-1.374) | 1.047<br>(0.784-1.399) |
| Ever had child live with<br>you | 1.344<br>(0.999-1.809) | 1.463*<br>(1.049-2.039) | 1.386*<br>(1.029-1.868) |
| *Sexual Orientation (Reference: Heterosexual)* |  |  |  |
| Gay/lesbian | 1.088<br>(0.246-4.809) | 2.163<br>(0.318-14.694) | 0.752<br>(0.198-2.854) |
| Bisexual | 1.838<br>(0.906-3.730) | 1.958<br>(0.789-4.860) | 1.576<br>(0.793-3.133) |
| *Household Income (Reference: High income - \$50k+)* | | | |
| Low income (<\$20k) | 1.041<br>(0.789-1.374) | 0.652*<br>(0.470-0.903) | 0.834<br>(0.636-1.094) |
| Middle income (\$20k to<br>< \$50k) | 1.249<br>(0.911-1.712) | 1.390<br>(0.942-2.051) | 1.373*<br>(1.001-1.883) |
| *Marital Status (Reference: Married)* |  |  |  |
| Divorced, separated, or<br>widowed | 2.233***<br>(1.610-3.098) | 1.250<br>(0.85-1.838) | 1.890***<br>(1.386-2.576) |
| Never married | 0.900<br>(0.668-1.210) | 0.711<br>(0.495-1.023) | 0.908<br>(0.676-1.221) |
| *Education (Reference: Primary education or some high school)* |  |  |  |
| High school diploma | 1.206<br>(0.834-1.743) | 0.999<br>(0.671-1.491) | 1.140<br>(0.791-1.643) |
| Vocational school | 1.352<br>(0.732-2.498) | 2.586*<br>(1.098-6.093) | 1.405<br>(0.764-2.582) |
| Some college | 1.359<br>(0.934-1.978) | 1.437<br>(0.944-2.187) | 1.313<br>(0.906-1.903) |
| College diploma or<br>above | 1.410<br>(0.905-2.196) | 1.726*<br>(1.032-2.888) | 1.367<br>(0.883-2.117) |
| *Health Status* |  |  |  |
| Perceived physical<br>health status | 0.903<br>(0.794-1.027) | 1.009<br>(0.872-1.169) | 0.921<br>(0.811-1.045) |
| Perceived mental health<br>status | 0.918<br>(0.811-1.039) | 1.056<br>(0.916-1.217) | 0.958<br>(0.848-1.084) |

\* p<0.05, \*\* p<0.01, \*\*\* p<0.001

### Formal supports

Closer analysis of specific types of support, namely police, medical, and therapy were conducted (Table 3). Compared to adult AIAN women survivors who are married, divorced, separated, or widowed adult AIAN women who experienced IPV in their lifetime have higher odds of seeking law enforcement support (OR=1.93, 95% CI=1.41 to 2.63) and higher odds of seeking therapy (OR=1.80, 95% CI=1.32 to 2.47). Those who never married are at lower odds of seeking medical support (OR=0.67, 95% CI=0.46 to 0.98) than those who are married. Compared to those who completed primary education or attended some high school, women who attended vocational school have odds as 1.98 times (CI= 1.00 to 3.91) as high of seeking medical care. Odds for seeking therapy are 1.86 times (95% CI=1.22 to 2.85) as high among those with some college experience and 1.99 times (95% CI=1.21 to 3.29) as high among those with a college diploma or higher. For every one-point increase in perceived mental health status, there are lower odds of seeking medical support (OR=0.78, 95% CI=0.68 to 0.91) and therapy (OR=0.83, 95% CI=0.73 to 0.95).

**Table 3.** Adjusted ORs of specific types of formal support

|  | Police | Medical | Therapy |
| --- | --- | --- | --- |
|  | Adjusted OR<br>(95% CI) | Adjusted OR<br>(95% CI) | Adjusted OR<br>(95% CI) |
| *Demographic* |  |  |  |
| Age 18-64 years | 1.003<br>(0.992-1.013) | 1.004<br>(0.993-1.016) | 0.997<br>(0.986-1.009) |
| Enrolled in tribe | 1.117<br>(0.826-1.512) | 1.019<br>(0.729-1.426) | 0.975<br>(0.717-1.327) |
| Ever had child<br>live with you | 1.210<br>(0.887-1.651) | 1.079<br>(0.765-1.520) | 0.934<br>(0.681-1.281) |
| *Sexual Orientation (Reference: Heterosexual)* |  |  |  |
| Gay/lesbian | 1.625<br>(0.352-7.511) | 1.194<br>(0.249-5.727) | 1.369<br>(0.352-5.326) |
| Bisexual | 0.898<br>(0.444-1.818) | 0.801<br>(0.364-1.762) | 1.611<br>(0.826-3.142) |
| *Household income (Reference: High income- \$50k+)* | | | |
| Low income<br>(<\$20k) | 1.113<br>(0.842-1.471) | 1.200<br>(0.878-1.641) | 0.843<br>(0.626-1.135) |
| Middle income<br>(\$20k to < \$50k) | 1.042<br>(0.747-1.453) | 1.071<br>(0.723-1.588) | 1.123<br>(0.793-1.590) |
| *Marital status (Reference: Married)* |  |  |  |
| Divorced,<br>separated, or<br>widowed | 1.927***<br>(1.411-2.632) | 1.372<br>(0.983-1.916) | 1.803***<br>(1.316-2.470) |
| Never married | 1.010<br>(0.741-1.377) | 0.670*<br>(0.460-0.975) | 0.764<br>(0.549-1.064) |
| *Education (Reference: Primary education or some high school)* |  |  |  |
| High school<br>diploma | 0.973<br>(0.666-1.421) | 1.314<br>(0.841-2.054) | 1.234<br>(0.799-1.903) |
| Vocational<br>school | 1.011<br>(0.542-1.888) | 1.979*<br>(1.002-3.908) | 1.827<br>(0.939-3.554) |
| Some college | 1.056<br>(0.723-1.540) | 1.526<br>(0.987-2.360) | 1.864**<br>(1.219-2.852) |
| College diploma<br>or above | 0.831<br>(0.530-1.305) | 1.047<br>(0.617-1.791) | 1.991**<br>(1.207-3.286) |
| *Health Status* |  |  |  |
| Perceived<br>physical health<br>status | 0.896<br>(0.784-1.023) | 1.028<br>(0.876-1.205) | 0.906<br>(0.786-1.043) |
| Perceived<br>mental health<br>status | 1.057<br>(0.930-1.202) | 0.783**<br>(0.676-0.906) | 0.831**<br>(0.726-0.953) |

\* p<0.05, \*\* p<0.01, \*\*\* p<0.001

### Informal supports

Three types of informal supports were examined: family, friend, and intimate partner. (Table 4) Compared to adult AIAN women survivors who are married, those who never married had lower odds of seeking family support (OR=0.66, 95% CI=0.48 to 0.91) and lower odds of seeking support from their current intimate partner (OR=0.46, 95% CI=0.32 to 0.66). Those who are divorced, separated, or widowed have lower odds (OR=0.56, 95% CI= 0.39 to 0.80) of seeking support from an intimate partner. Compared to adult AIAN women with high household income, those in low-income households have lower odds of seeking support from an intimate partner (OR=0.71, 95% CI=0.52 to 0.98). Compared to respondents who completed primary school or attended some high school, odds are 2.20 times (95% CI=1.12 to 4.30) as high for seeking support from an intimate partner among those who attended vocational school and 1.77 times (95% CI=1.05 to 2.97) as high among those who have a college diploma or higher. For every one-year increase in age, odds of seeking support from an intimate partner decrease (OR=0.98, 95% CI=0.97 to 0.99). Compared to those who are not enrolled in a tribe, the odds of adult AIAN women survivors seeking support from a friend is 8.33 times as high (95% CI=4.81 to 14.42).

**Table 4.** Adjusted ORs of specific types of informal support

|  | Family | Friend | Intimate Partner |
| --- | --- | --- | --- |
|  | Adjusted OR<br>(95% CI) | Adjusted OR<br>(95% CI) | Adjusted OR<br>(95% CI) |
| *Demographic* |  |  |  |
| Age 18-64 years | 0.995<br>(0.984-1.006) | 0.990<br>(0.964-1.018) | 0.980**<br>(0.968-0.992) |
| Enrolled in tribe | 1.018<br>(0.749-1.382) | 8.330***<br>(4.814-14.415) | 0.816<br>(0.590-1.127) |
| Ever had child<br>live with you | 1.169<br>(0.861-1.587) | 0.942<br>(0.454-1.954) | 0.869<br>(0.621-1.217) |
| *Sexual Orientation (Reference: Heterosexual)* |  |  |  |
| Gay/lesbian | 0.941<br>(0.211-4.195) | 0.324<br>(0.037-2.857) | 1.135<br>(0.228-5.642) |
| Bisexual | 0.709<br>(0.366-1.377) | 2.170<br>(0.434-10.865) | 1.660<br>(0.807-3.414) |
| *Household income (Reference: High income- \$50k+)* | | | |
| Low income<br>(<\$20k) | 0.791<br>(0.595-1.052) | 1.416<br>(0.697-2.876) | 0.714*<br>(0.519-0.982) |
| Middle income<br>(\$20k to < \$50k) | 1.213<br>(0.868-1.695) | 1.273<br>(0.636-2.550) | 0.727<br>(0.516-1.023) |
| *Marital status (Reference: Married)* |  |  |  |
| Divorced,<br>separated, or<br>widowed | 0.776<br>(0.564-1.067) | 1.476<br>(0.708-3.075) | 0.559**<br>(0.390-0.801) |
| Never married | 0.662*<br>(0.482-0.909) | 1.320<br>(0.590-2.951) | 0.462***<br>(0.324-0.656) |

| *Education (Reference: Primary education or some high school)* |  |  |  |
| --- | --- | --- | --- |
| High school diploma | 0.740<br>(0.505-1.085) | 0.527<br>(0.207-1.344) | 1.181<br>(0.743-1.876) |
| Vocational school | 1.208<br>(0.628-2.322) | 0.670<br>(0.152-2.960) | 2.197*<br>(1.123-4.298) |
| Some college | 0.921<br>(0.628-1.350) | 0.696<br>(0.251-1.934) | 1.509<br>(0.966-2.359) |
| College diploma or above | 0.947<br>(0.601-1.493) | 0.798<br>(0.266-2.400) | 1.767*<br>(1.053-2.966) |
| *Health Status* |  |  |  |
| Perceived physical health status | 1.001<br>(0.876-1.145) | 1.048<br>(0.784-1.400) | 0.890<br>(0.762-1.039) |
| Perceived mental health status | 1.127<br>(0.992-1.280) | 1.253<br>(0.946-1.660) | 1.113<br>(0.959-1.291) |

\* p<0.05, \*\* p<0.01, \*\*\* p<0.001

## Discussion

Formal support alone is not often sought out. However, among the formal supports, there are higher odds of seeking therapy. Formal supports seem to have higher odds of being utilized when informal supports are also sought, particularly among divorced, separated, or widowed women, those who ever had a child live with them, and those in middle income households. The odds of seeking police were only significant high with divorced, separated, or widowed women. They also have significantly higher odds of seeking therapy. Police is not often regarded as a support as there is general mistrust towards law enforcement and the criminal justice system among AIAN communities. Attitudes towards law enforcement are that there are delays and that they are insufficient in providing protection for the victim [9]. There may also be issues with trusting the overall criminal justice system as the perpetrator may not be adequately held accountable, or there may be a lack of consistency in how the case is handled. While the results of seeking police are unsurprising, this study initiates further inquiry on why divorced, separated, or widowed women have higher odds of seeking support from the police. Police may become involved if the abuser is an ex-partner as studies indicate that perpetrators are the most violent when women try to separate from their abusive partners [21]. While there were no significant odds of seeking specific types of support among those who ever had a child live with them, it is important to note that other types of formal support that were not included in this analysis, such as legal and housing supports.

Those who attended vocational school have significantly higher odds of seeking medical support. Those who are never married or with better perceived mental health status had lower odds of seeking medical support. Seeking health services are intertwined with fears of being victim-blamed and being subjected to negative assumptions [10]. There are significantly higher odds of seeking therapy among those who are divorced, separated, or widowed, those with some college experience, and those with a college diploma or higher. However, it is unclear whether this is associated with reasons such as access to campus resources, increased awareness of therapy services, or reduced stigma regarding mental healthcare.

Seeking only informal support appears to be more common, particularly among those who showed significant higher odds, such as those who ever had a child live with them, those who attended vocational school, and those with a college diploma or higher. However, closer examination reveals no higher odds of seeking family support, and those who were never married had significantly lower odds of seeking family support. Compared to those who completed primary school or attended some high school those went to vocational school or have a college diploma or above have significantly higher odds of seeking support from an intimate partner. Factors such as being divorced, separated, or widowed, being never married, low household income, and age have significantly lower odds of seeking support from a friend. Tribal enrollment had significantly higher odds (OR=8.33, 95% CI=4.81 to 14.42) of seeking support from a friend. The majority (81%) of respondents reported being enrolled, which may contribute to the result.

This study provides ample opportunity to inquire further about support-seeking behavior among adult AIAN women survivors of IPV. For instance, the odds of seeking police support were higher (though not significant) for most of the variables, except for those who are bisexual, those with a high school diploma and perceived physical health status. From Table 1, 45% of respondents sought the police but 77% of those who sought the police considered it to be helpful. This prompts future research on how survivors define “helpful” and whether their needs were qualitatively met through police support. Odds ratio analyses show that attending vocational school or having a college diploma have higher odds for seeking support from a friend, but lower odds for multiple variables. Only 29% of respondents report seeking support from an intimate partner, but 94% of those perceive it to be helpful. Intimate partners could potentially be a supportive resource for survivors, but education on how to engage with survivors and support them may be in need-especially for those who are divorced, separated, or widowed, those who never married, those with low household income, and those who are older in age. While 69% of respondents sought support from a friend and of those 97% perceived it to be helpful, the only significant relationship was observed with tribal enrollment (OR=8.33, 95% CI=4.81 to 14.42). Those living on reservations are likely to be enrolled. However, there are many urban AIANs who are also enrolled. Further research is needed to understand the relationship between tribal enrollment and seeking support from a friend. In addition, AIANs who are aware of their tribal affiliations but do not have enrollment for any reason may present similar or different relationship to seeking support.

## Limitations

In this study, odds ratios were calculated to understand the general relationships between various individual, community, relational, and systemic factors with support-seeking behavior. However, there is much that is yet to be understood regarding AIAN women’s experiences with seeking support. The dataset focuses on the individual’s behavior in a limited time-frame relative to the specific experiences of victimization reported and does not adequately capture the societal and cultural barriers that the women may experience. Follow-up studies that qualitatively inquire the nuances of support seeking behavior may provide valuable insights.

### Public health implications

Systemic barriers to seeking care include lack of funding for shelters or health centers, and fear that medical and legal personnel are not sympathetic towards Natives due to misperceptions, stereotypes, and prejudice. There are also systemic delays in health care services due to the perception that the Bureau of Indian Affairs or the Indian Health Service provides all needed assistance. Further, there is lack of coordination among law enforcement operating under a fragmented criminal justice system. This is unfortunately not unique to the United States, as it is stated to be a critical issue even in Canada (Razack, 2016).

Given the significant high risk of IPV among adult AIAN women, there is an urgent need for adequate survivor support. While there are various avenues in which survivors may seek support, there is limited understanding as to how these decisions are made and what factors play significant roles in this process. This study provides insight into how certain factors inform the odds of seeking support. The helpfulness of a specific type of support does not provide clarity on the decision-making processes of support-seeking behavior and the nuances that come with access, cultural barriers, and victim-blaming. However, this study is still valuable in distinguishing factors related to support-seeking and observing general relationships. Focused and intentional public health research on support for AIAN women survivors is needed to further understand how programs and policies can be improved.

## Data Availability

Data used for this study is from the National Intimate Partner and Sexual Violence Survey (NISVS) 2010 (ICPSR 36140). It is restricted data that can be obtained through ICPSR with appropriate IRB approval.

## Acknowledgements

I thank Monique Castro, LMFT from the Indigenous Circle of Wellness and Dr. Carrie Johnson from Seven Generations Child & Family Services for their support. I also thank Dr. Jangho Yoon for providing guidance for the conceptual framework and analysis.

## Footnotes

2 Moving forward, “Native American,” “Native,” “American Indian,” “AIAN,” and “Indigenous” are used interchangeably depending on the context.

